# Stepping it up: A Longitudinal Assessment of Stair Negotiation Among ACL Reconstruction Patients Outside the Laboratory

**DOI:** 10.1101/2024.05.16.24307484

**Authors:** Tomer Yona, Bezalel Peskin, Arielle Fischer

## Abstract

**Introduction:** Anterior Cruciate Ligament Reconstruction (ACLR) is crucial for knee stability in ACL-injured individuals and for resuming pre-injury physical activities. Despite advancements, achieving symmetrical movement patterns during rehabilitation, particularly in stair negotiation, poses challenges. This study examines lower limb kinematics during stair negotiation at various rehabilitation stages post-ACLR, employing inertial measurement units (IMUs) and Statistical Parametric Mapping (SPM) for in-depth analysis outside the laboratory.

**Methods:** This cohort study longitudinally assessed stair ambulation kinematics in patients aged 18-40, three (n=26) and five months (n=18) post-ACLR, using IMUs to track sagittal plane movement during stair ascent and descent. The participants ambulated on a flight of 20 stairs outside the laboratory.

**Results:** At three months post-ACLR, the injured knee was less flexed compared to the contralateral knee during stair ascent (mean difference = -11.3°, CI [-14.4, -8.1], p<.001) and descent (mean difference = -6.3°, CI [-10.2, -2.4], p=.002). SPM analysis identified clusters where the injured knee showed decreased flexion at 0-35% and 87-99% of the stair ascent cycle (p<.005). By five months, flexion differences during ascent improved (mean difference = -4.7°, CI [-8.1, -1.4], p=.008), but significant asymmetry persisted, with decreased flexion at 10-32% of the cycle during ascent and 20-29% during descent (p<.017). Improvements between three and five months were observed in knee flexion during ascent (mean increase = 6.1°, p<.001) and descent (mean increase = 9.3°, p=.004). Ankle and hip joint movements also exhibited persistent asymmetries, with minimal improvements over time.

**Conclusions:** Persistent lower limb kinematic asymmetries remain five months post-ACLR during stair ascent and descent.

## Introduction

Annually, approximately 1.5 million active individuals face a considerable challenge: the rupture of their Anterior Cruciate Ligament (ACL), a crucial stabilizer in the knee.^1,2^ This injury not only sidelines them from the activities they love but also initiates a long and uncertain journey towards recovery, leading to immediate disability and long-term consequences, including lower quality of life, movement asymmetries, and the early onset of knee osteoarthritis.^3^

ACL reconstruction (ACLR) is an accepted surgery to restore knee stability and function and reduce secondary knee injuries. However, the rehabilitation process and the return to pre-injury level activities remain challenging, with patients often showing altered kinematics and kinetics during various activities.^4,5^ Specifically, stair negotiation, a seemingly mundane activity, becomes a goal toward independence for individuals after an ACLR.

Biomechanically, stair negotiation is particularly relevant for post-ACLR assessment as it requires greater knee flexion angles, challenging the knee’s stability. Previous studies have highlighted that individuals with ACLR demonstrate significant biomechanical deviations during stair negotiation, indicating compensatory strategies or persistent deficits in knee function.^5–8^ However, these studies are limited to laboratory settings using a low number of steps (2-5 stairs), which may not accurately represent real-world movements.

Assessing stair biomechanics, particularly during the mid-stage of rehabilitation, is vital, as this period is important for achieving symmetrical movement patterns.^9^ By five months, individuals often transition to more demanding activities, making it a pivotal time to assess whether early rehabilitation efforts have successfully restored functional biomechanics or if compensatory strategies persist.

Recently, 1-dimensional Statistical Parametric Mapping (Spm1d) was introduced to the biomechanics community, allowing continuous data to be analyzed and provide a more comprehensive view of the movement.^10^ While Spm1d can potentially identify subtle changes in movement patterns that traditional discrete point analysis may overlook, a recent review found that it is not yet commonly used to assess stair negotiation among patients after an ACLR.^11^ Additionally, using inertial measurement units (IMUs) enables the assessment of kinematics in environments where patients live outside the confines of a laboratory setting.

By conducting this study outside the laboratory environment using a real flight of stairs and spm1d together with discrete point analysis, we aim to address those gaps and offer an ecological approach to stair biomechanics post-ACLR.

The aims of this study are (a) to describe the knee kinematics during stair ambulation at three and five months post-ACLR and (b) to evaluate the changes in the knee kinematics between three and five months post-ACLR. A secondary aim was to describe the kinematics and kinematic changes of the hip and ankle during stair ambulation at three and five months post-ACLR.

## Methods

A cohort study design, approved by the Rambam health care campus Helsinki committee (0089-21-RMB), was employed to longitudinally assess the kinematics of stair ambulation among patients’ changes during their mid-stage rehabilitation after an ACLR. This study is part of a randomized clinical trial (NCT05001594). We recruited patients aged 18-40, three months after an ACLR and without previous lower limb injuries. All the participants were recruited from Rambam Health Care Campus, Haifa, Israel, from 2020 to 2023 and were operated by one of five surgeons. A single physiotherapist assessed the patient’s routine check-up at the hospital.

Each participant ascended and descended a flight of 20 stairs with a rise of 17cm and a run of 30cm. This was repeated three times for 60 steps at their comfortable, self-selected speed. A full cycle was defined from weight acceptance to the following weight acceptance of the same leg (Figure 1). Each trial’s first and last two steps were removed from the analysis.

**Figure 1.**
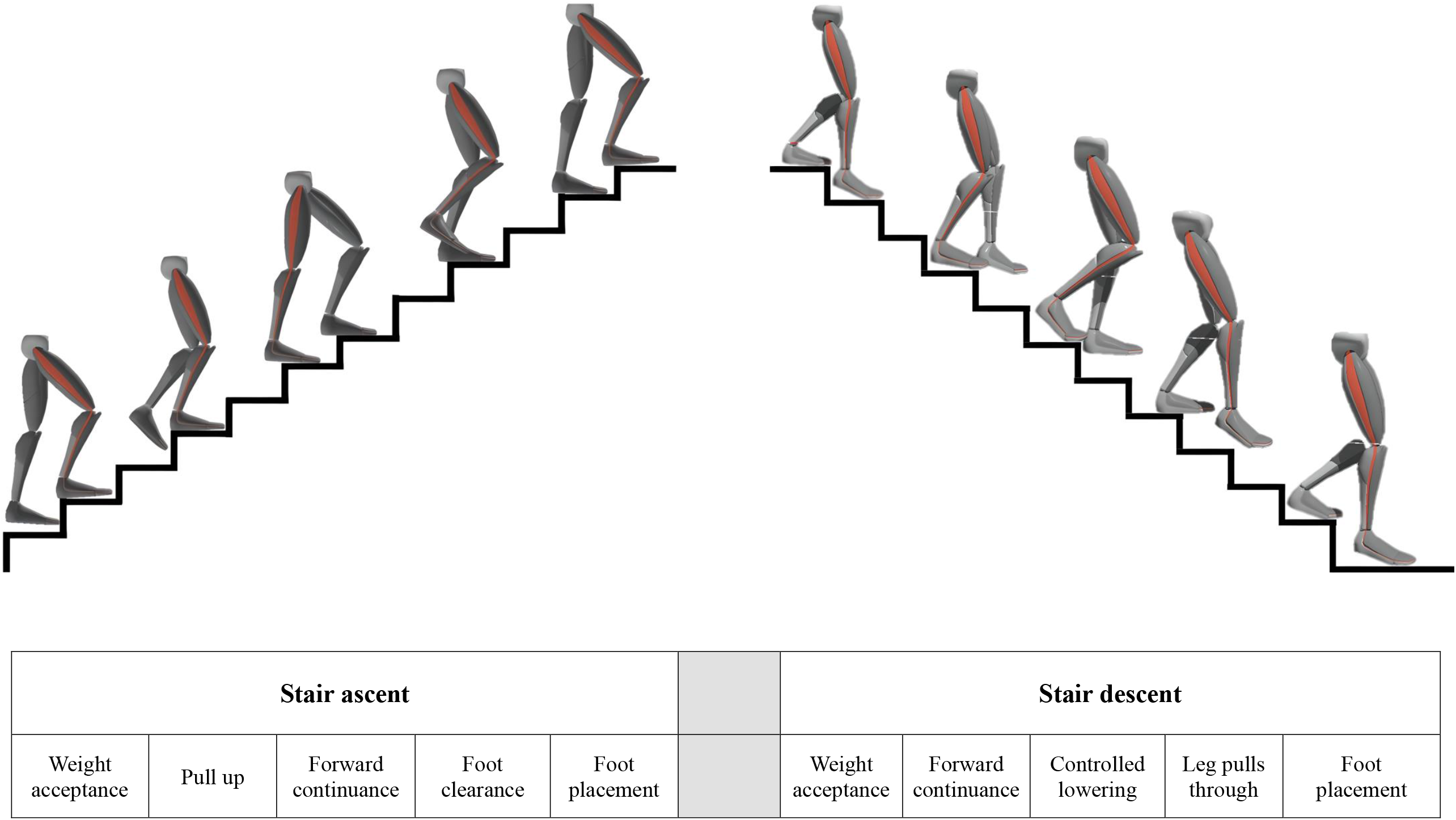
Stair ambulation movement cycle The different parts of the movement cycle of stair ambulation

**Figure 2.**
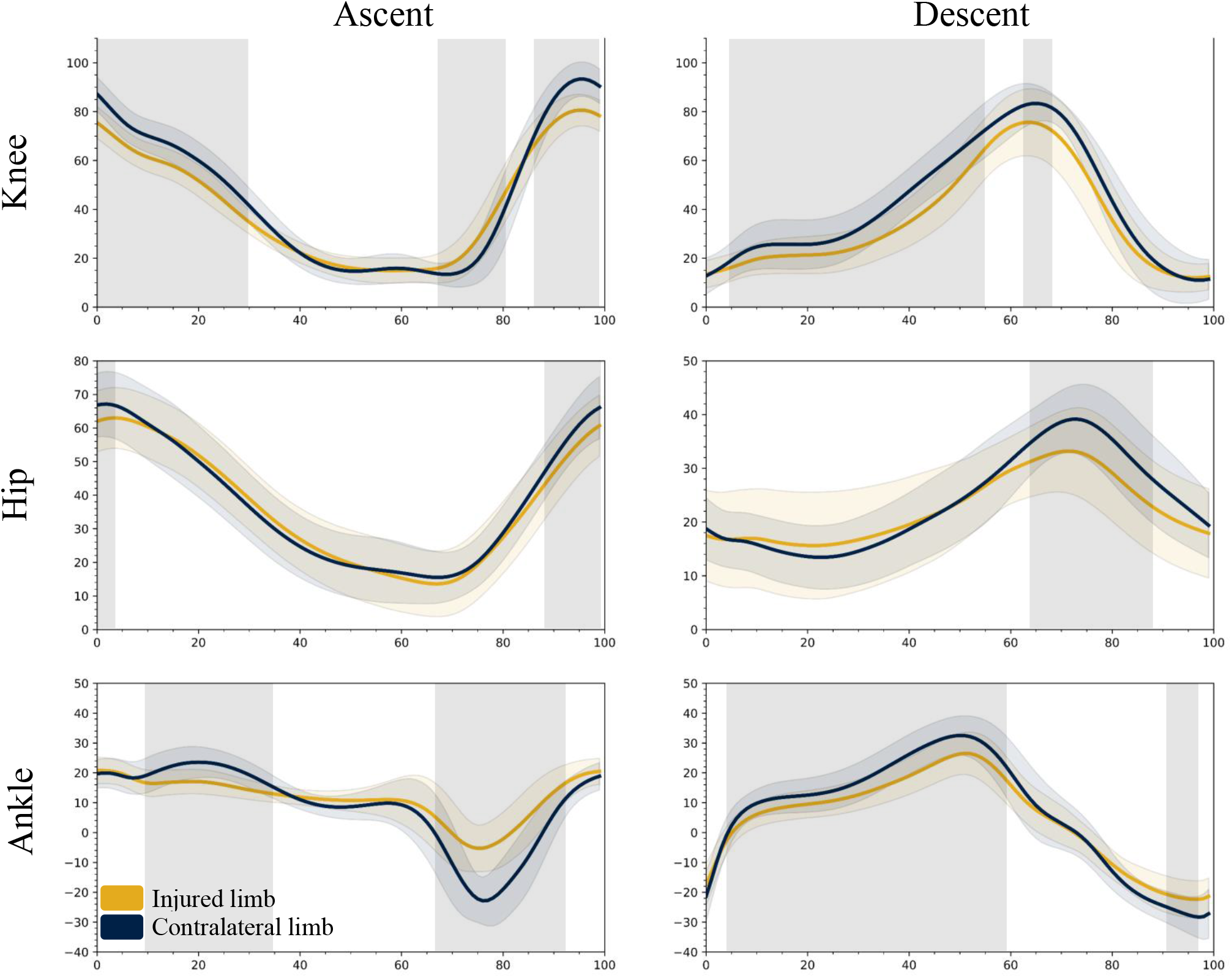
The average sagittal movements during the full stair ascent and descent cycle comparing the injured and contralateral limb at three months post-ACLR (n=26). One-Dimensional Statistical Parametric Mapping (spm1d) Paired t-test analysis. Boxed shaded grey area = statistically significant difference between the injured and contralateral leg.

The participants were allowed to rest as needed between each trial. The lower limb sagittal kinematics were collected using seven IMUs located on the pelvis, thighs, shanks, and feet (XSENS Awinda, Movella). This inertial motion capture system has been validated and assessed for reliability, especially for sagittal plane movements.^12^ As few studies have used IMUs to assess stair ambulation, we recruited a control group of healthy controls, matched by height and weight, to establish a baseline for normal stair ambulation patterns using IMUs.

The sample size calculation is based on a previous study that assessed knee kinematic asymmetries between the injured and contralateral knees six months post-ACLR. The study reported 81.8±7.4 peak flexion for the injured knee and 85.7±5.7 for the contralateral knee.^13^ Using G*Power’s (version 3.1.9.6) f test for matched pairs, with a significance criterion of α = .05 and power = .80, the total sample size needed was 20 participants.^14^

The normality of the data was assessed using the Shapiro-Wilk test. Descriptive statistics are reported as mean ± standard deviation. Differences between the limbs were evaluated using the paired sample t-test with Cohen’s d effect size [95% confidence interval]. Linear mixed models were used to assess changes between the time points and to account for missing data. A post hoc sensitivity analysis was conducted using multiple imputations (100 computed linear regression imputations) techniques.^15^ Spm1d paired t-test was used to determine differences between the full kinematic waves of each joint.^10^

We performed all the discrete points analyses using IBM SPSS (Version 29). spm1d paired sample t-tests were used to assess the differences between the injured and contralateral limb and between the times (spm1D v0.4.18, http://www.spm1d.org). The alpha level was set at 0.05 for all statistical tests.

## Results

Of the 26 patients recruited three months post-ACLR, 18 attended the five-month check-up. The participants’ demographic data are presented in Table 1. For the healthy controls, there was no statistically significant differences between the any of the joints of the left and right limbs during stair ambulation (Appendix 1).

**Table 1.** Demographics of the participants.

|  | **ACLR**<br>**Participants**<br>**(n=26)** | **Healthy**<br>**Participants**<br>**(n=16)** | **p** |
| --- | --- | --- | --- |
| **Age (years)** | 23.5 ± 5.7 | 29.7 ± 5.0 | <.001 |
| **Sex (%)** |  |  |  |
| Males | 21 (77.8%) | 12 (75.0%) | .835 |
| Females | 6 (22.2%) | 4 (25.0%) |  |
| **Height (cm)** | 177 ± 0.1 | 167 ± 0.1 | .131 |
| **Weight (kg)** | 75.6 ± 11.7 | 64.6 ± 14.8 | .163 |
| **Injured Leg (%)** |  |  |  |
| Left | 18 (45%) | N/A | N/A |
| Right | 22 (55%) |  |  |
| **Time between injury and reconstruction (days)** | 248 ± 165 | N/A | N/A |
| **Graft (%)** |  |  |  |
| Hamstrings | 17 (63.0%) | N/A | N/A |
| BTB | 5 (18.5%) |  |  |
| Quadriceps | 4 (14.8%) |  |  |
| Allograft | 1 (3.7%) |  |  |

The independent samples t-test was used to compare nominal data. The $\chi^2$ test was used to compare categorical data.

### Three months post-ACLR

#### Full movement analysis (SPM1d)

As seen in Figure 1, at three months post-ACLR, the mean knee flexion angle of the injured leg was lower than the contralateral knee at 0-35%, 87-99%, and higher at 68-81% of the cycle, as seen by three supra-threshold clusters exceeding the critical threshold of t[1,25]=3.15, p<.005. During stair descent, the injured knee was less flexed compared to the contralateral knee at 5-55% and 63-68% of the cycle, as seen by two supra-threshold clusters exceeding the critical threshold of t[1,25]=3.14, p<.005

The mean hip flexion of the injured leg was lower than the angle of the contralateral hip at 0-3% and 88-99%, as seen by two supra-threshold clusters exceeding the critical threshold of t[1,25]=3.05, p<.005. During stair descent, the injured hip was less flexed compared to the contralateral knee at 64-88% of the cycle, as seen by a single supra-threshold cluster exceeding the critical threshold of t[1,25]=3.03, p=.001

The mean ankle dorsiflexion angle of the injured leg was lower than the angle of the contralateral ankle at 10-35% and 68-93%, as seen by two supra-threshold clusters exceeding the critical threshold of t[1,25]=3.20, p<.005. During stair descent, the injured hip was less flexed compared to the contralateral knee at 4-59% and 91-98% of the cycle, as seen by two supra-threshold clusters exceeding the critical threshold of t[1,25]=3.23, p<.005

### Discrete point analysis

Table 2 shows that, compared to the contralateral limb, the injured knee was less flexed at the maximum point while ascending (mean difference = -11.3, CI [-14.4, -8.1], p<.001) and descending stairs (mean difference = -6.3, CI [-10.2, -2.4], p=.002). There was no statistically significant difference in the knee minimum points.

**Table 2.** Description of the hip, knee, and ankle movements in the sagittal plane while **ascending and descending** stairs three and five months after an ACL reconstruction surgery.

| Joint | Angle | Three months post-surgery<br>(n=26) |  |  |  | Five months post-surgery<br>(n=18) |  |  |  |
| --- | --- | --- | --- | --- | --- | --- | --- | --- | --- |
|  |  | Involved | Contralateral | p | ES (CI) | Involved | Contralateral | p | ES (CI) |
| Stair Ascent |  |  |  |  |  |  |  |  |  |
| Hip | Minimum | 12.8 ± 9.3 | 15.3 ± 7.4 | .029 | **-0.45 [-0.85, -0.04]\*** | 12.2 ± 9.1 | 15.4 ± 8.5 | .029 | **-0.56 [-1.05 - -0.05]\*** |
|  | Maximum | 64.0 ± 8.3 | 67.4 ± 8.8 | .003 | **-0.64 [-1.06, -0.21]\*** | 66.3 ± 9.1 | 68.3 ± 9.7 | .048 | **-0.50 [-0.98 - -0.01]\*** |
| Knee | Minimum | 13.4 ± 4.6 | 11.0 ± 4.1 | .006 | 0.38 [-0.01, 0.78] | 11.1 ± 4.8 | 10.8 ± 4.7 | .857 | 0.04 [-0.42, 0.50] |
|  | Maximum | 82.7 ± 7.2 | 94.0 ± 6.1 | <.001 | **-1.43 [-1.98, -0.87]\*** | 87.6 ± 3.9 | 92.4 ± 5.6 | .008 | **-0.71 [-1.22, -0.18]\*** |
| Ankle | Minimum | -9.2 ± 6.7 | -25.1 ± 7.6 | <.001 | **2.01 [1.33, 2.68]\*** | -14.0 ± 5.6 | -24.6 ± 7.5 | <.001 | **1.71 [0.96 – 2.43]\*** |
|  | Maximum | 22.0 ± 4.2 | 24.9 ± 3.2 | .006 | **-0.58 [-0.99, -0.16]\*** | 20.8 ± 3.5 | 24.9 ± 2.9 | <.001 | **-1.01 [-1.58 - -0.43]\*** |
| Stair Descent |  |  |  |  |  |  |  |  |  |
| Hip | Minimum | 13.0 ± 8.5 | 12.0 ± 6.8 | .381 | 0.17 [-0.21, .056] | 10.4 ± 7.7 | 12.4 ± 5.9 | .140 | -0.36 [-0.83, 0.11] |
|  | Maximum | 33.9 ± 8.3 | 39.4 ± 7.0 | <.001 | **-0.95 [-1.41, -0.48]\*** | 35.1 ± 7.1 | 37.3 ± 7.9 | .071 | -0.45 [-0.93, 0.03] |
| Knee | Minimum | 12.1 ± 5.1 | 10.2 ± 5.6 | .113 | 0.32 [-0.07, 0.71] | 11.1 ± 3.4 | 11.7 ± 3.7 | .598 | -0.12 [-0.58, 0.33] |
|  | Maximum | 79.9 ± 11.4 | 86.2 ± 5.9 | .002 | **-0.66 [-1.07, -0.22]\*** | 88.3 ± 5.1 | 87.7 ± 5.6 | .605 | 0.12 [-0.33, 0.59] |
| Ankle | Minimum | -24.2 ± 5.5 | -29.5 ± 5.8 | <.001 | **0.83 [0.37, 1.21]\*** | -25.8 ± 4.1 | -28.1 ± 4.0 | .059 | 0.47 [-0.01, 0.96] |
|  | Maximum | 27.7 ± 6.5 | 34.6 ± 6.7 | <.001 | **-1.16 [-1.65, -0.65]\*** | 31.2 ± 4.7 | 36.1 ± 5.0 | .002 | **-0.85 [-1.38, -0.30]\*** |

For between limbs comparison: Paired sample t-test, Cohen's d effect size [95% confidence interval]. \* Denotes a statistically significant difference between the injured and the contralateral limb.

While ascending stairs, the injured hip was less flexed at both the minimum (mean difference = -2.4, CI [-4.7, -0.2], p=.029) and maximum point (mean difference = -3.4, CI [-5.6, -1.2], p=.003) and only at the maximum point while descending stairs (mean difference = -5.5, CI [-7.9, -3.1], p<.001).

Lastly, during stair ascent, the injured ankle was less dorsiflexed at the minimum point (mean difference = - 15.8, CI [-12.6, -19.0], p<.001) and less plantarflexed at the maximum point (mean difference = -2.8, CI [-4.8, -0.9], p=.006). Similar results were found for the minimum (mean difference = -5.3, CI [-2.7, -7.9], p<.001) and the maximum points (mean difference = -6.9, CI [-9.3, -4.5], p<.001) at during descending stairs.

### Five months post-ACLR

#### Full movement analysis

As seen in Figure 3, the mean knee flexion angle of the injured leg was lower than the contralateral knee at 10-32% of the cycle, as seen by a single supra-threshold cluster exceeding the critical threshold of t[1,17]=3.25, p=.001. Similarly, during stair descent, the injured knee was less flexed than the contralateral knee at 20-29% of the cycle, as seen by a single supra-threshold cluster exceeding the critical threshold of t[1,17]=3.36, p=.017.

**Figure 3.**
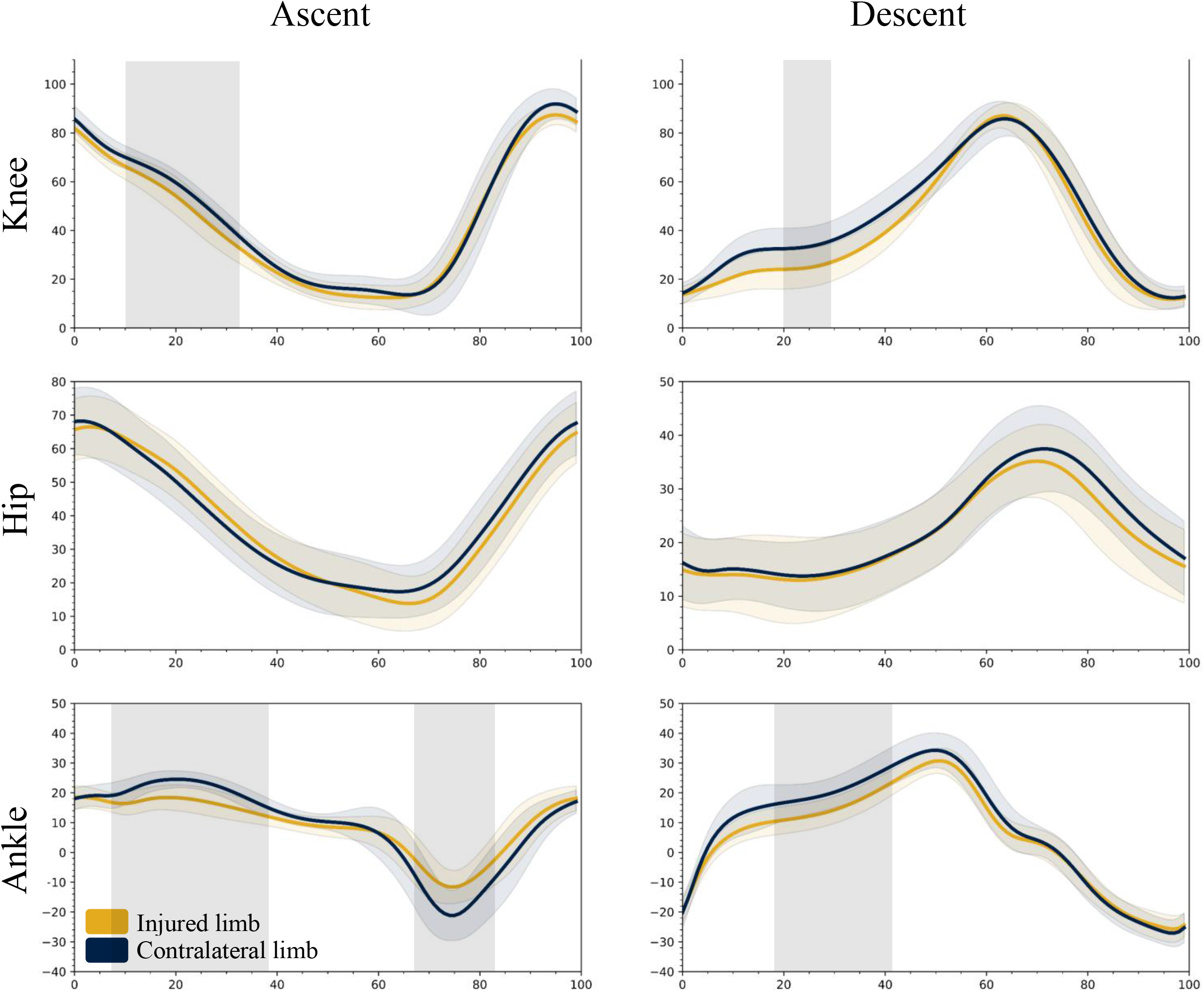
The average sagittal movements during the full stair ascent and descent cycle comparing the injured and contralateral limb at five months post-ACLR (n=18). One-Dimensional Statistical Parametric Mapping (spm1d) Paired t-test analysis. Boxed shaded grey area = statistically significant difference between the injured and contralateral leg.

No differences were found between the injured and contralateral hips during ascending and descending.

The mean ankle dorsiflexion angle of the injured leg was lower than the angle of the contralateral ankle at 8-38% and less plantarflexed at 67-83%, as seen by two supra-threshold clusters exceeding the critical threshold of t[1,17]=3.46, p<.005. During stair descent, the injured ankle was less dorsiflexed at 18-41% of the cycle, as seen by a single supra-threshold cluster exceeding the critical threshold of t[1,17]=3.42, p<.001.

#### Discrete points analysis

Table 2 shows that, compared to the contralateral limb, the injured knee was less flexed at the maximum point only when ascending stairs (mean difference = -4.7, CI [-8.1, -1.4], p=.008).

While ascending stairs, the injured hip was less flexed at both the minimum (mean difference = -3.1, CI [-6.0, -0.3], p=.029) and maximum point (mean difference = -2.0, CI [-4.0, -0.2], p=.048). There were no significant differences at the hip joint while descending stairs.

During stairs ascending, the injured ankle was less dorsiflexed at the minimum point (mean difference = 10.6, CI [7.5, 13.6], p<.001) and less plantarflexed at the maximum point (mean difference = -4.1, CI [-6.2, -2.1], p<.001). Lastly, during stair descending, the injured ankle was less plantarflexed at the maximum point (mean difference = -4.9, CI [-7.8, -2.0], p<.001).

### Between time differences

#### Full movement analysis

During stair ascent, the knee mean flexion angle increased between three months and five months post ACLR at 0-12% and 83-99% of the cycle, as seen by two supra-threshold clusters exceeding the critical threshold of t[1,16]=3.38, p<.005. A similar pattern was observed during stair descent at 60-68% of the cycle, as seen by a single supra-threshold cluster exceeding the critical threshold of t[1,16]=3.35, p=.027

For the hip angles, no differences were found between three and five months post ACLR during stair ascending and descending.

During stair ascent, the ankle angle increased from three months to five months post ACLR at 74-80% and 85-95% of the cycle, as seen by two supra-threshold clusters exceeding the critical threshold of t[1,16]=3.41, p<.005. No differences were found during the stair descent.

#### Discrete points analysis

The discrete point analysis is presented in Table 3.

**Table 3.** Differences between three- and five-months post ACLR in the hip, knee, and ankle movements in the sagittal plane while ascending and descending stairs.

| Joint | Angle | Stair Ascend |  |  |  | Stair Descend |  |  |  |
| --- | --- | --- | --- | --- | --- | --- | --- | --- | --- |
|  |  | Three Months | Five Months | Difference | p-value | Three Months | Five Months | Difference | p-value |
| Hip | Minimum | 12.6 ± 1.9 | 12.6 ± 2.1 | ?N/A | .996 | 12.7 ± 1.7 | 9.9 ± 1.9 | -2.7 ± 1.8 | .144 |
|  | Maximum | 63.4 ± 1.7 | 66.6 ± 2.0 | 3.2 ± 1.9 | .112 | 33.4 ± 1.6 | 34.9 ± 2.0 | 1.5 ± 2.2 | .505 |
| Knee | Minimum | 13.5 ± 0.9 | 11.0 ± 1.1 | -2.5 ± 0.9 | **.019\*** | 12.0 ± 0.9 | 11.6 ± 1.0 | -0.3 ± 1.0 | .716 |
|  | Maximum | 82.7 ± 1.3 | 88.8 ± 1.4 | 6.1 ± 1.2 | **<.001\*** | 79.5 ± 1.9 | 88.8 ± 2.4 | 9.3 ± 2.9 | **.004\*** |
| Ankle | Minimum | -9.5 ± 1.2 | -14.3 ± 1.5 | -4.8 ± 1.4 | **.004\*** | -24.3 ± 1.0 | -26.3 ± 1.1 | -1.9 ± 0.9 | .057 |
|  | Maximum | 22.0 ± 0.8 | 20.8 ± 0.9 | -1.1 ± 1.1 | .319 | 27.3 ± 1.2 | 31.3 ± 1.4 | 3.9 ± 1.5 | **.018\*** |

Linear mixed models assessments of between times differences of the lower limb kinematics (mean ± standard error). \* Denotes a statistically significant difference between timepoints by post-hoc pairwise comparison.

#### Sensitivity analysis

Sensitivity analysis was performed using three different techniques. The analysis yielded similar results for the a-priori-defined primary outcome, the knee-related analysis. However, the knee minimum angle at stair ascent at the three-month point significantly differed between the injured and contralateral limbs.

For the hip joint, differences were found in all parameters but the hip maximum angle at stair descent in the between-times comparison. All the other hip parameters were statistically significant in this analysis. For the ankle joint during stair descent, the minimum angle at five months post and the between-times analysis is statistically significant under this analysis.

## Discussion

The main aim of our study was to describe the lower limb kinematics of patients three and five months after ACLR while ascending and descending a flight of 20 stairs. We add new evidence to the literature by longitudinally assessing stair ambulation during mid-stage ACLR rehabilitation, using IMUs to collect ecologically valid data, and using spm1d together with discrete point analysis to describe the findings. We found that at three months post-ACLR, significant deviations exist in the injured leg, spanning the hip, knee, and ankle joints, compared to the contralateral leg. However, noticeable improvements are seen from three to five months post-ACLR, indicating progress in the rehabilitation but highlighting persistent kinematic compensation from the ankle joint during ascending and descending stairs.

The inconsistencies of stair ambulation assessments in previous studies make it challenging to compare them directly with our results. Many have chosen to analyze only the weight-bearing phase, possibly because the cycle segmentation was related to a force plate, making the segmentation easy as the start and end of the cycle are visible using the force vector from the plate.^13,16,17^ Most studies were observational studies at a single time point or with multiple time points with different participants.^17,18^ Next, only a handful of studies assessed the hip and ankle.^13,17^ Lastly, previous studies focused on late assessment after an ACLR (6 months-23 years), with a single study by Markström et al., who evaluated a group early after their reconstruction.^17^

Looking at a group of patients 2.7 months after ACLR, Markström et al. evaluated the kinematics of the weight acceptance phase during stair descent. Contrary to their findings of no differences in knee movement in the sagittal plane, we found lower flexion angles during stair descent, including in the stance phase. Next, while they did not find differences in the hip movements, our reported differences were not in the stance phase, explaining the differences between the studies. Lastly, our results are on par with those of Markström et al. Regarding the ankle asymmetries, both report lower plantarflexion at the stance phase.^17^

The improvement between time points agrees with others who assessed how the hip and ankle asymmetries improve over time.^17^ Further, others have found that asymmetries in the knee might stay for 20 years after the ACLR.^8,16^ Our study suggests that despite improvements, patients do not fully achieve kinematic symmetry at five months post-op at any of the lower limb joints and extends previous observations to a real-world setting, emphasizing the ecological validity of our findings.

Our study underscores the persistent kinematic compensations at three and five months post-ACLR, highlighting the need for tailored rehabilitation strategies that address asymmetries. By using IMUs, clinicians can gain more insights into their patients’ progress and functional limitations outside the controlled environment of their clinic. This can lead to a more dynamic rehabilitation process that adapts to the patient’s needs over time, potentially leading to improved outcomes.

Our study is not without limitations. The sample size, although sufficient for detecting significant differences, may limit the generalizability of our findings. Additionally, the use of IMUs, while increasing the ecological validity of our findings, is less accurate than optoelectrical camera systems. Therefore, we conducted a sensitivity analysis, increasing the robustness of the results. Additionally, the multiple comparisons in our study increase the chance for Type I errors, and we defined the knee kinematics in the sagittal plane as our primary outcome, as it is directly affected by the ACLR. Next, some of the secondary outcomes are different in the sensitivity analysis, which might reduce the robustness of our results regarding the hip and ankle, requiring further studies focusing on those joints. Lastly, we did not assess differences between graft types, sexes, and time from injury to surgery. Therefore, future longitudinal studies should follow up beyond five months post-ACLR, focus on the ankle and hip as the primary outcomes, and assess sex and graft differences.

## Conclusion

This study underscores the complexities of achieving kinematic symmetry after ACLR during stair negotiation. Despite the observed improvements between the three and five-month postoperative time points, kinematic asymmetries persist in the knee, hip, and ankle.

## Data Availability

All data produced in the present study are available upon reasonable request to the authors

---

**Appendix 1.**
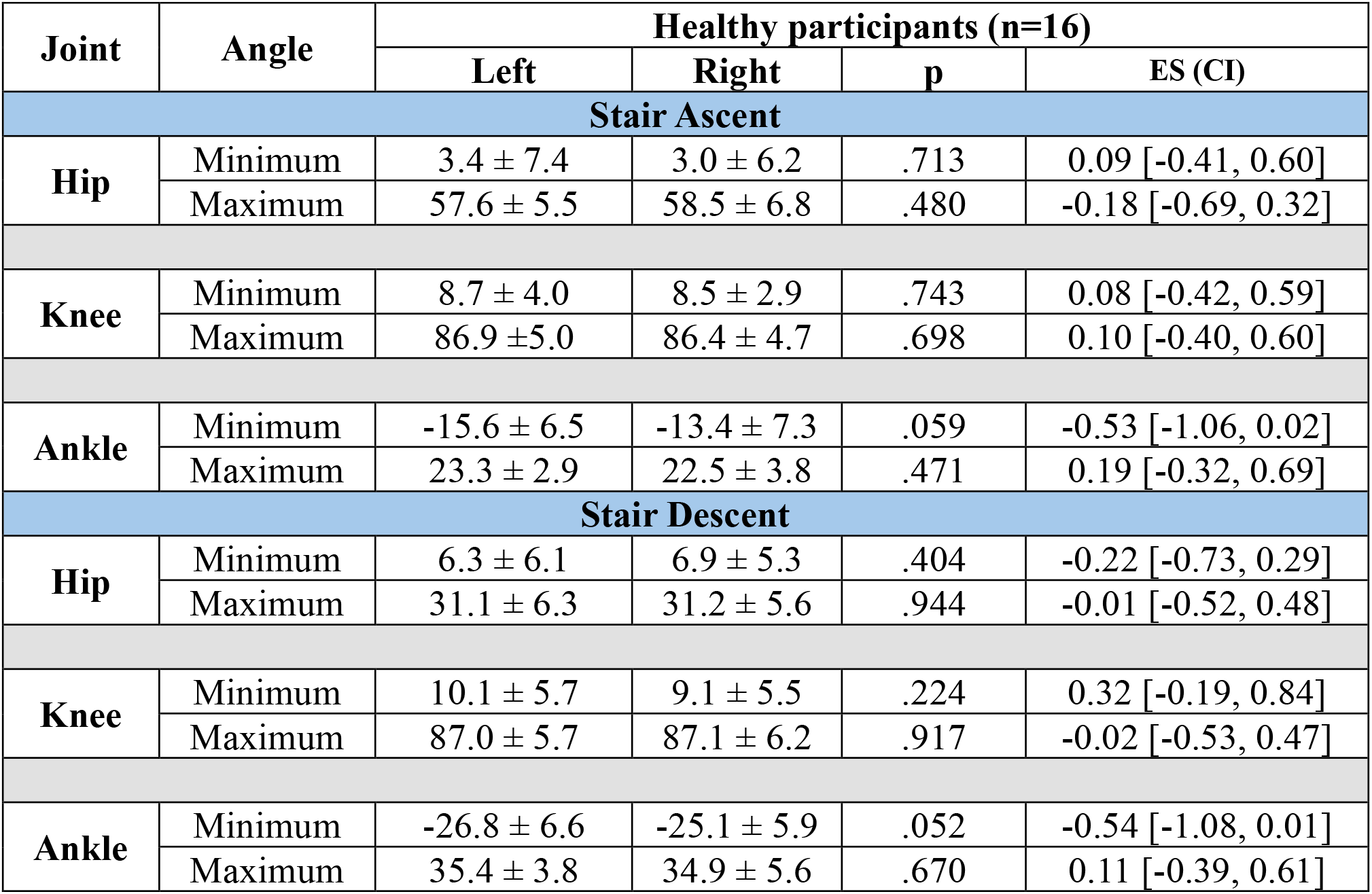
Descriptive statistics of the lower limb sagittal kinematics of the healthy participants.

| Joint | Angle | Healthy participants (n=16) |  |  |  |
| --- | --- | --- | --- | --- | --- |
|  |  | Left | Right | p | ES (CI) |
| Stair Ascent |  |  |  |  |  |
| Hip | Minimum | 3.4 ± 7.4 | 3.0 ± 6.2 | .713 | 0.09 [-0.41, 0.60] |
|  | Maximum | 57.6 ± 5.5 | 58.5 ± 6.8 | .480 | -0.18 [-0.69, 0.32] |
| Knee | Minimum | 8.7 ± 4.0 | 8.5 ± 2.9 | .743 | 0.08 [-0.42, 0.59] |
|  | Maximum | 86.9 ±5.0 | 86.4 ± 4.7 | .698 | 0.10 [-0.40, 0.60] |
| Ankle | Minimum | -15.6 ± 6.5 | -13.4 ± 7.3 | .059 | -0.53 [-1.06, 0.02] |
|  | Maximum | 23.3 ± 2.9 | 22.5 ± 3.8 | .471 | 0.19 [-0.32, 0.69] |
| Stair Descent |  |  |  |  |  |
| Hip | Minimum | 6.3 ± 6.1 | 6.9 ± 5.3 | .404 | -0.22 [-0.73, 0.29] |
|  | Maximum | 31.1 ± 6.3 | 31.2 ± 5.6 | .944 | -0.01 [-0.52, 0.48] |
| Knee | Minimum | 10.1 ± 5.7 | 9.1 ± 5.5 | .224 | 0.32 [-0.19, 0.84] |
|  | Maximum | 87.0 ± 5.7 | 87.1 ± 6.2 | .917 | -0.02 [-0.53, 0.47] |
| Ankle | Minimum | -26.8 ± 6.6 | -25.1 ± 5.9 | .052 | -0.54 [-1.08, 0.01] |
|  | Maximum | 35.4 ± 3.8 | 34.9 ± 5.6 | .670 | 0.11 [-0.39, 0.61] |

